# Physical Activity, Symptoms, Quality of Life and Exercise Program Preferences in People with Chronic Lymphocytic Leukemia

**DOI:** 10.1101/2025.02.10.25321744

**Authors:** Ellie E. Miles, Jennifer L. Nicol, Hatti Fowler, Amelia Roberts, Andrew T. Hulton, Caitlin Jeary, Renata Walewska, Sunil Iyengar, Erik D. Hanson, CLL Support Association, Andrea Sitlinger, David B. Bartlett

## Abstract

**Background:** Chronic lymphocytic leukaemia (CLL) has a heterogeneous lifelong course. Most patients experience significant symptoms which negatively impact their quality of life (QoL). Although physical activity and exercise may help manage symptoms, it is unclear what disease-related factors drive physical inactivity in people with CLL.

**Methods:** This study explored physical activity in CLL and assessed relationships in treatment stage, symptoms, quality of life, and preferences for physical activity using an online questionnaire.

**Results:** 128 individuals with CLL (66M/62F) completed the questionnaire. Treated CLL (N=55) exhibited worse QoL (p=0.018) and lower engagement in physical activity (p=0.045) compared to treatment naïve (N=73). Both groups reported high fatigue (∼77%) and insomnia (∼55%) which were associated with less likelihood of being physically active. Physically active participants reported better QoL (p=0.020), physical functioning (P=0.003) and role functioning (p=0.020) as well as lower levels of fatigue (p=0.036), pain (p=0.017) and symptom burden (p=0.026). Although 79% of respondents wanted to engage in exercise programs for their CLL, 70% reported never receiving exercise guidance from their healthcare professionals.

**Conclusion:** Findings highlight a significant need for interventions to increase physical activity in people with CLL. Furthermore, there is considerable interest from the CLL community in receiving exercise guidance.

## Introduction

Chronic lymphocytic leukaemia (CLL) is the most common adult blood cancer in the Western world, with approximately 3,800 new cases diagnosed annually and over 20,000 prevalent cases in the United Kingdom (1, 2). CLL primarily affects males (63% of cases) and is largely a disease of older adults, with an average age of diagnosis over 70 years old (3). However, over the past decade, the incidence of CLL has increased by 25% among individuals aged 50-70 years, and is projected to keep rising as diagnostic tools are improved (1). People with CLL experience a complex disease journey characterised by an increased risk for infections and secondary cancers, with high symptom burden and comorbidities that reduce their quality of life (QoL). Central to this are cancer-associated and premature age-associated reductions in physical function and physiologic reserve (4, 5). Reduced functional performance is associated with poor survival, more infections and further reductions in QoL (4, 6). Further compounding this complexity are the additional symptoms and reductions in QoL of those patients on active monitoring (i.e., dynamic monitoring, treatment naïve or watch and wait), which differs, in parts, from those receiving treatment (7, 8). For those on active monitoring, the period between diagnosis and treatment is several years, and in some cases, treatment is never required and is associated with distress, anxiety, social isolation and fatigue (7, 8). Similarly, for those on treatment, there are treatment-related symptoms (e.g., nausea, diarrhoea, bleeding, fatigue) that compound the years of reduced QoL experienced whilst awaiting treatment. Therefore, there is a growing need to understand the relationships between symptoms, QoL and physical function to determine whether interventions will reduce CLL-specific symptoms and improve QoL (9).

Although increasing physical activity levels and exercise exposure are recommended for managing symptoms and improving QoL in several solid cancers (10), it is unclear whether this is generalisable to CLL. Compared to solid cancers, patients with haematologic malignancies have lower physical function, higher levels of fatigue and an increased risk of frailty (11). Approximately 60-70% of older adults with CLL are classified as either pre-frail or frail, a striking contrast to the 15-30% frailty observed in the general older population (12, 13). Physical activity and exercise-based interventions can often reduce frailty by improving physical function in both clinical and community settings (14). Furthermore, in haematological malignancies, being more physically active is associated with a better QoL (15). Recently, small pilot studies of exercise interventions in patients with CLL suggest that exercise training with behaviour change strategies to increase physical activity can increase physical fitness and physical function, lower symptom burden and improve overall QoL (16–18). Despite the potential benefits of physical activity and enhanced physiological fitness for individuals with CLL, physical activity levels are 30-40% lower than those of healthy individuals in the same age group (5). It is unclear what factors drive this physical inactivity phenotype and whether there is an opportunity for healthcare professionals to intervene with lifestyle approaches. Therefore, it is crucial to identify and address factors influencing physical activity and exercise participation in this population while also gaining a deeper understanding of how physical activity impacts QoL in individuals with CLL.

This study aimed to investigate for the first time the relationships between physical activity, QoL, CLL symptoms, and comorbidities among individuals with CLL at different stages of treatment. In addition, we aimed to assess the beliefs and preferences related to physical activity and exercise advice and delivery to understand if there is an opportunity for healthcare professionals to promote exercise interventions.

## Methods

### Study Population

We recruited individuals diagnosed with CLL through the CLL Support Association UK. This patient-led charity supports people with CLL or small lymphocytic leukaemia (SLL), families, and supporters. Recruitment was through word of mouth, online promotion, and clinic referrals. Information on participants’ diagnoses was self-reported and collected through an initial screening questionnaire. People were eligible if they had a confirmed diagnosis of CLL or SLL of any treatment status, were aged 18 years or over, could understand the written English language, had access to a computer/laptop/tablet/phone and could provide written informed consent. Participants were provided with details of the aims and requirements for the study through the participant information sheet before the consent process. Those eligible who completed the consent form then gained access to the questionnaire via Qualtrics (Qualtrics LLC, USA). The University of Surrey Ethics Committee reviewed the protocol and gave a favourable ethical opinion (FHMS 21-22 261 EGA).

### Survey Instrument Development

We collected data from this cross-sectional observation study using an online version (Qualtrics LLC, USA) of a 183-part questionnaire separated into nine sections (Supplementary Figure 1).

#### Section 1 (Items 1-8) – Personal Information

Participant demographics, current living situation, mode of transport to medical consultations, and highest level of education.

#### Section 2 (Items 9-33) – Self-reported health- and disease-related characteristics

Date of diagnosis, current treatment stage; Treatment-Naïve, individuals who have not received treatment and are currently going through a period of observation (Active Monitoring); In Treatment, individuals who are currently undergoing active treatment for CLL; Post-Treatment, individuals who have previously undergone treatment for CLL but are currently in remission (2), current and previous treatments, current healthcare; private or National Health Service (NHS), and severity of CLL-related symptoms experienced in the past month (classified as ‘not at all’, ‘slightly’, ‘moderately’, ‘severely’ and ‘overwhelmingly’).

#### Section 3 (Items 34-41) – Self-reported physical activity levels

Participants were presented with two separate validated and reliable physical activity questionnaires. The Stanford Brief Activity Survey (SBAS) (19, 20) and the Godin Leisure Time Physical Activity Questionnaire (Godin LTPAQ) (21). The Godin LTPAQ is commonly used to classify individuals into active and insufficiently active categories, with a moderate correlation between Godin LTPAQ and a pedometer step count in leukaemia and other cancer survivors having been observed (22, 23). The Godin LTPAQ was used to understand current physical activity levels; however, the questionnaire was also adapted to gauge an understanding of physical activity levels before diagnosis. Godin LTPAQ physical activity categories were determined by moderate and vigorous intensity units per week where ≥24 units were classified as “Active”, 14-23 units as “Moderately Active”, and <14 units as “Insufficiently Active”. However, when categorising into whether participants met physical activity guidelines or not, the time spent in moderate and vigorous activity was summed, with the minutes in vigorous activity weighted by two to account for greater intensity. Those who scored 150 minutes per week or greater were categorised as meeting physical activity guidelines (24, 25).

#### Section 4 (Items 42-75) – Interest and advice on physical activity

Participants were questioned on the physical activity advice and guidance they had received from their healthcare professionals, their interest in participating in an exercise programme and their preferences regarding the delivery of an exercise programme. Responses were collected using a 5-point Likert scale ranging from 1 (strongly disagree/very unimportant) to 5 (strongly agree/very important). The questions used in this section were based on previous research into how individuals with Multiple Myeloma, a blood cancer with similar symptoms to CLL (e.g. fatigue, muscle weakness), view exercise and their preferences for participating in physical activities (24).

#### Section 5 (Items 76-121) – Self-reported quality of life (QoL)

Participants were asked questions from the English version of the European Organisation for Research and Treatment of Cancer Quality of Life Questionnaires - Core30 (EORTC QLQ-C30 v3) (26, 27), including the extension for CLL (EORTC QLQ-CLL17) (28). All scales range in scores from 0 to 100; a high score represents a higher (better) level of functioning or a higher (worse) level of symptoms. (29).

#### Section 6 (Items 122-151) – CLL Comorbidities Index

Participants were presented with questions on their present and past medical status to understand their comorbidity burden. Questions covered cardiac, vascular, haematological, respiratory, EENT (eyes, ear, nose, throat, larynx), upper/lower GI, hepatic, pancreatic, renal, genitourinary, musculoskeletal, neurological, endocrine-metabolic, and psychiatric/Behavioural conditions. Each condition was then scored using the Cumulative Illness Rating Scales (30) adapted to CLL (31). The sum of the system scores creates an overall comorbidity index.

#### Section 7 (Items 152-176) - Dietary Analysis

To evaluate diet quality, participants responded to questions from a short-form food frequency questionnaire (SFFFQ) (32). A dietary quality score (DQS) was derived from their intake of fruits, vegetables, oily fish, non-milk extrinsic sugars, and fats, which are considered key indicators of a healthy diet. For each component, scores ranged from 1 to 3, with a score of 3 indicating adherence to UK dietary guidelines for that food group. The DQS ranged from a minimum of 5 to a maximum of 15, representing the optimal intake of these foods. This questionnaire has been validated by comparing it with a comprehensive food frequency questionnaire (FFQ), showing a significant agreement in DQS between the SFFFQ and FFQ within a UK population (32).

#### Section 8 (Items 177-183) - Additional Self-reported Health and Disease Characteristics

This section included information that not all participants may have had access to or know, which could have posed a barrier to completing the questionnaire. Questions included recent medical test results (i.e., Complete blood count results and Cytogenic Results (2)) and anaemia diagnosis. It was clear to the participants that these questions were optional.

### Statistical Analysis

We conducted statistical analyses using SPSS v29.0 (IBM, USA). We report the characteristics of our study population using descriptive statistics such as means ± standard deviation (SD) or median and interquartile range (IQR), depending on data normality determined by the Kolmogorov-Smirnov test. For data presentation and interpretation, we also present frequencies/proportions. We used Pearson Chi-square tests to assess categorical variables. Where data were normally distributed, we used T-tests and ANOVAs to compare groups. Where data were not normally distributed, nonparametric Mann-Whitney (continuous variables), Kruskal-Wallis (more than 2 groups) and the Wilcox Signed Rank test for paired samples were used. When appropriate, effect sizes were assessed using the rank-biserial correlation coefficient (r) and post hoc analysis was carried out using Dunn’s pairwise test. Significance values reported from the Dunn’s pairwise test have been adjusted by the Bonferroni correction for multiple tests.

Symptom intensities were described as: “not at all” (1)/“slightly” (2)/“moderately” (3)/“significantly” (4)/”overwhelmingly” (5) and collapsed into three categories: not at all (response 1), slightly (response 2), and moderately or greater (responses 3, 4, and 5). Preferences in exercise program responses were also collapsed into three categories to determine proportions. “strongly agree” (1)/”agree” (2)/”neutral” (3)/”disagree” (4)/”strongly disagree (5) was collapsed into agree (response 1 & 2), neutral (response 3), and disagree (response 4 & 5), while “very important” (1)/”important” (2)/”neutral” (3)/”unimportant” (4)/”very unimportant (5) was collapsed into important (response 1 & 2), neutral (response 3), and unimportant (response 4 & 5). Due to reduced numbers, we collapsed treatment status into two categories: Treatment-Naïve (TN) and Treated (TRE).

Multivariable logistic regression analyses were conducted to assess the factors associated with being physically active. Potential predictors were independently modelled depending on their classification as either clinical/demographic (e.g. age, BMI, years with CLL, years treated for CLL, comorbidity index), symptoms or QoL indices. Physical activity exposure/levels were the dependent variable and dichotomised into either insufficiently active or moderate/highly active. We present adjusted odds ratios with 95% confidence intervals (95% CI). All statistical tests were two-tailed, with an alpha level of ≤0.05 set for statistical significance, while an alpha level of ≤0.10 was considered a trend.

## Results

### Participant Characteristics

One hundred and twenty-eight people with confirmed CLL/SLL [66M/62F: mean age 67±9.1 years (range 38 - 91 years)] with treatment naïve [N=73 (57%)] or having received treatment [N=55 (43%)] completed the questionnaires. We present the socio-demographic and clinical characteristics in Table 1. Briefly, both TN and TRE groups of participants were a similar age (p=0.145), while treated participants reported being diagnosed with CLL for around 2.8 years longer (Z = −3.737, p<.001, *r* = 0.334). As expected, more treated participants reported a higher Binet stage than treatment naïve (p<0.001). We observed a trend for 50% more comorbidities in the treated group (Z = −1.839, p = 0.066, *r* = 0.163). In both groups, most participants were of white ethnicity, accounting for 100% of treatment naïve and 90.9% of treated individuals. A significant proportion of participants in both groups had a high level of education, with 80.9% and 69.2% of treatment naïve and treated individuals, respectively, holding at least an undergraduate degree or higher.

**Table 1.**
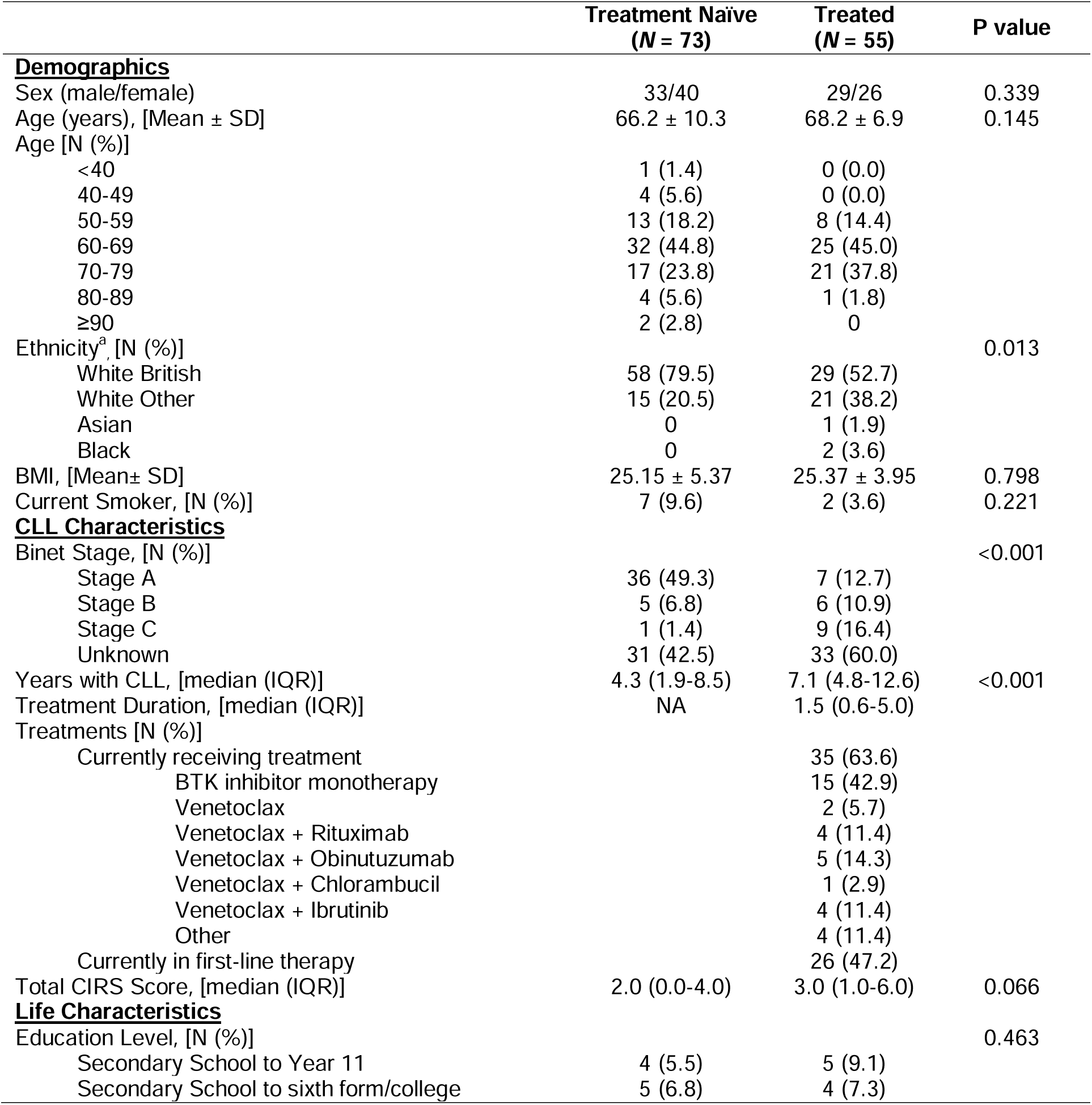
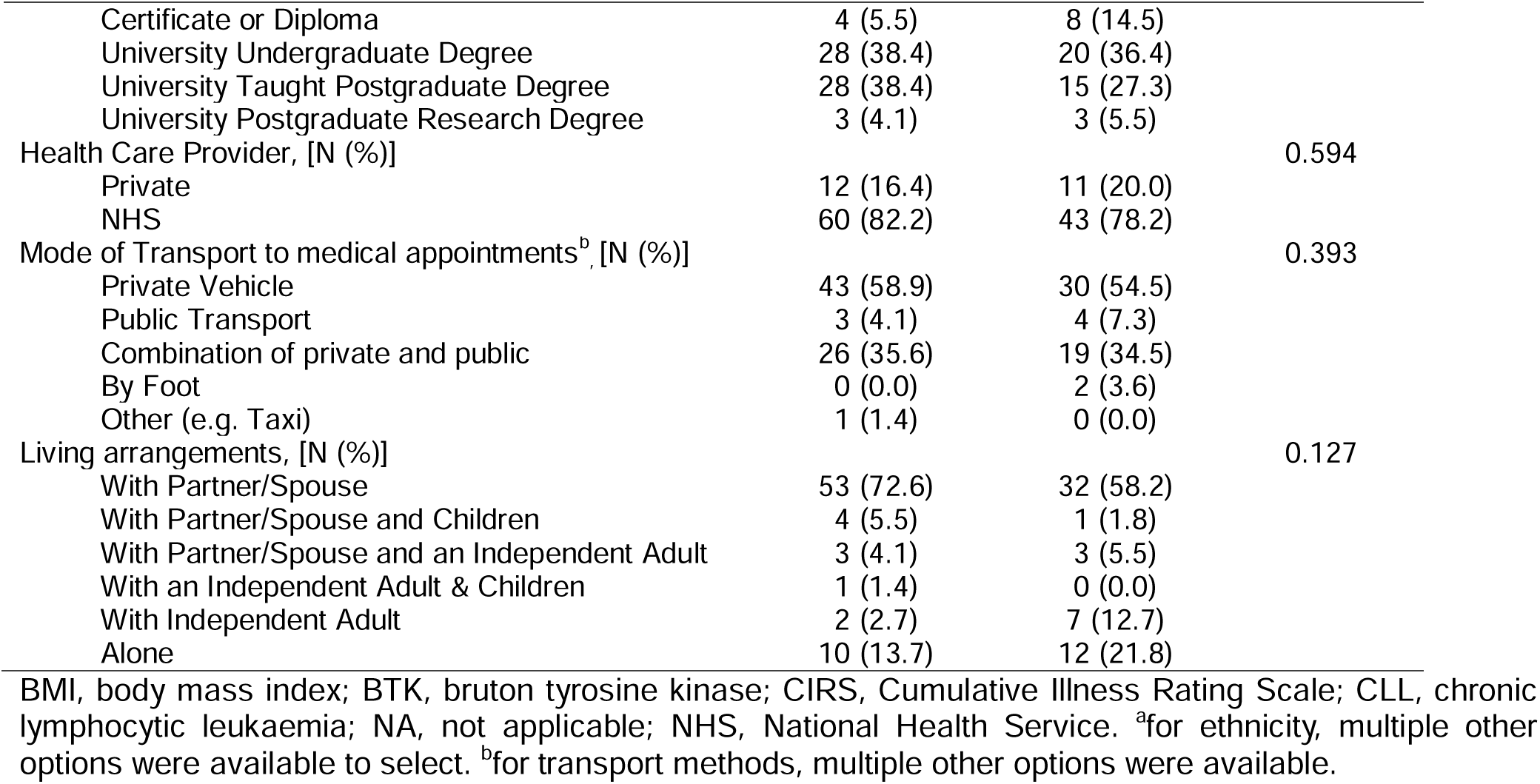
Socio-demographic and clinical characteristics by treatment status.

### Impact of Treatment Status on QoL

Figure 1 shows that when compared to treatment naïve participants, treated participants had a lower QoL as measured by global health status (Z = −2.740, p = 0.006, *r* = −0.250) and lower social functioning (Z = - 2.360, p = 0.018, *r* = −0.220). Additionally, treated participants showed a trend towards lower role functioning (Z = −1.710, p = 0.087, *r* = −0.160). No significant differences were found between groups on other functional scales.

**Figure 1:**
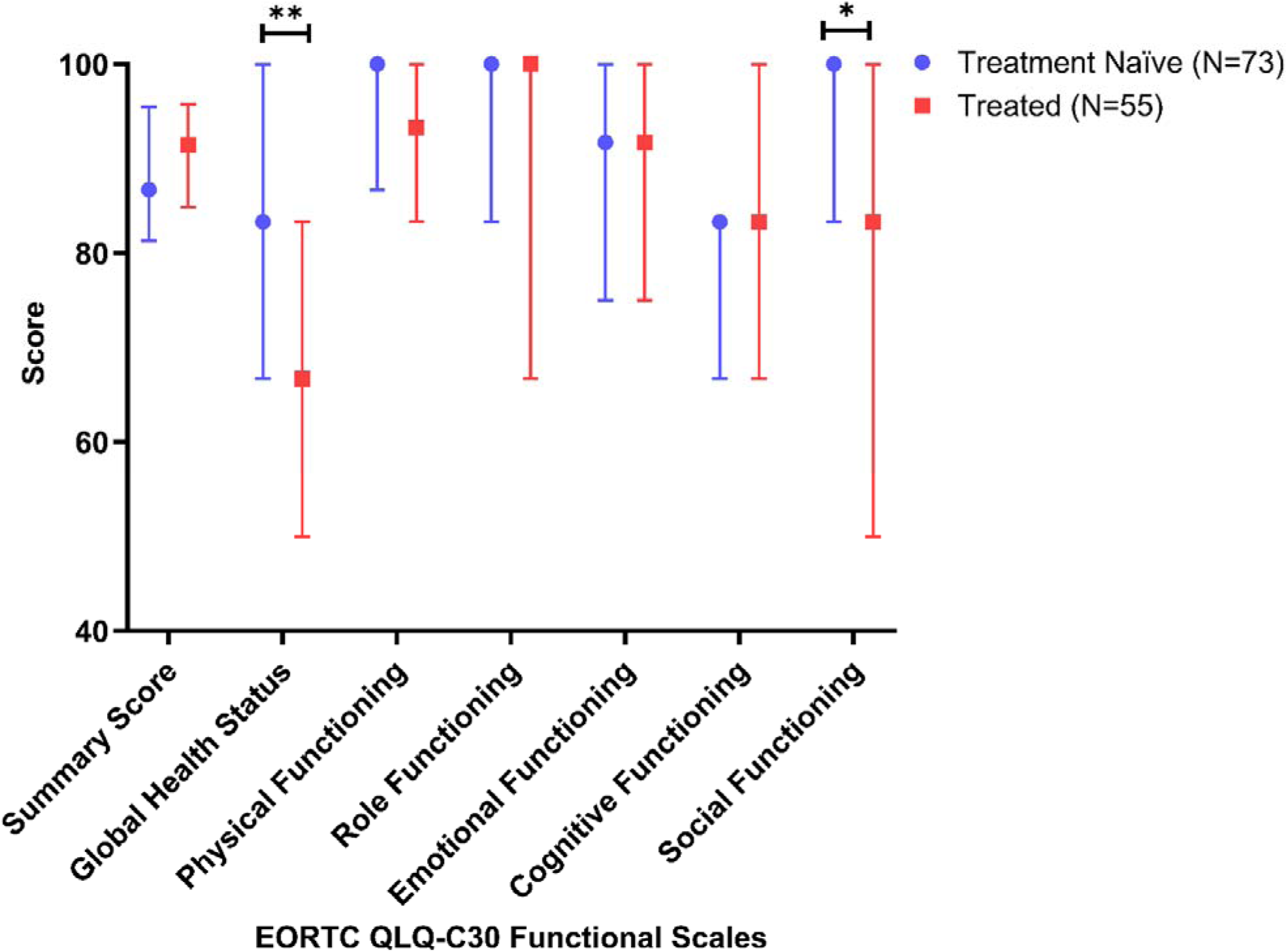
Comparison of global health status and functional scales of quality of life using EORTC QLQ-30 v3 Questionnaire in treatment naïve CLL participants compared to treated CLL. All the scales range in scores from 0 to 100; a high score represents a higher (better) level of functioning. Values are median and interquartile ranges. *p-value < 0.05, **p-value < 0.01

### Impact of Treatment Status on Symptoms

The frequency and intensity of CLL symptoms participants experienced in the month before completing the survey (Section 2 - <u>Self-reported health- and disease-related characteristics)</u> are presented in Figure 2A (treatment naïve) and Figure 2B (treated). Most respondents experienced at least one symptom at a moderate or greater intensity (65.3%), with 92.9% of individuals experiencing at least one symptom to a slight degree. Overall, fatigue was the most reported symptom in both groups, with around 77% of all participants experiencing fatigue. Although both groups reported similar feelings of fatigue, there was a trend for approximately 15% more treated participants experiencing higher levels of fatigue (p = 0.081). Similarly, more than 50% of participants in both groups reported insomnia, while 58.2% of treated participants reported stress. All other symptoms were reported in less than 50% of participants. When comparing other symptom profiles between groups, 25.6% more treatment naïve participants reported lymphadenopathy (p = 0.007) and twice as many treatment naïve participants reported unexpected weight loss (p = 0.012), while 10.2% more treated participants reported weakness (p = 0.044).

**Figure 2:**
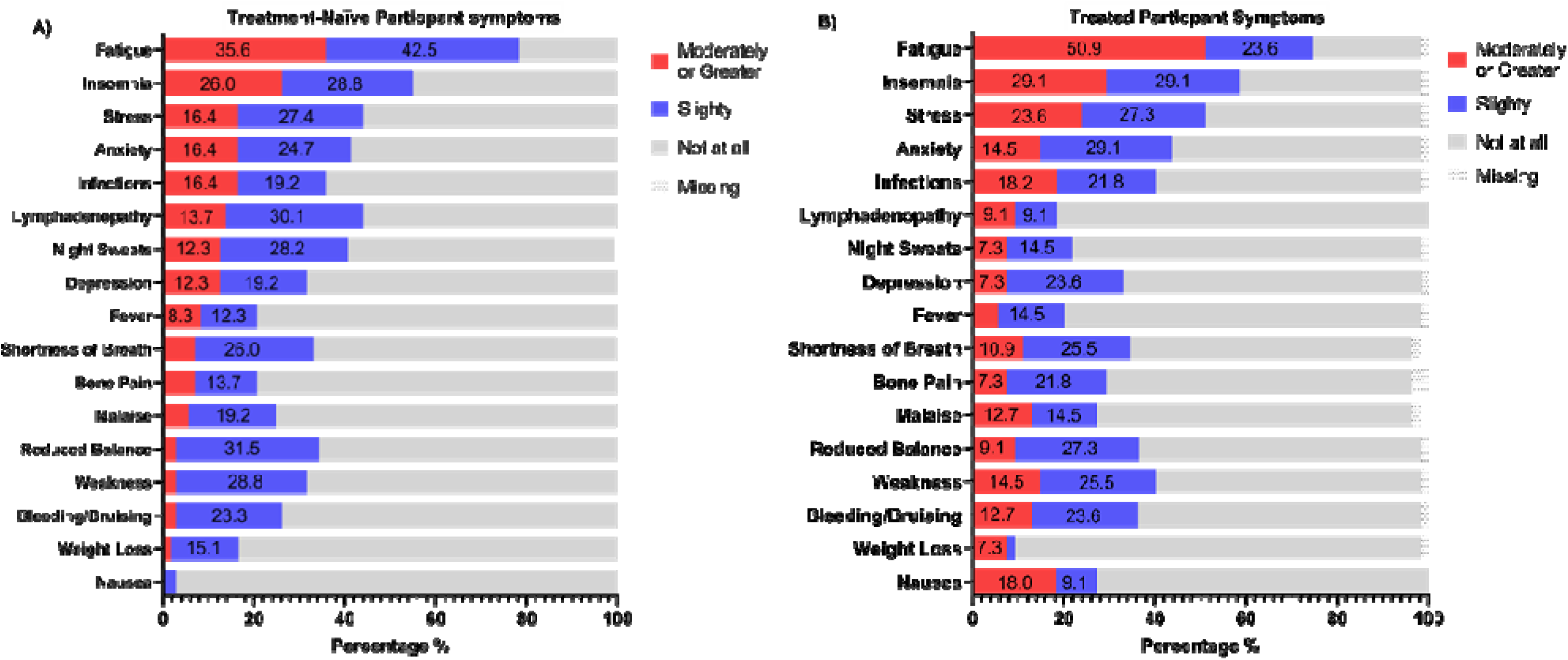
Intensity of symptoms over the previous month experienced by people with treatment naïve CLL (A: N=77) and peoples who have been treated for CLL (B: N=55).

When analysing EORTC QLQ-C30 and EORTC QLQ-CLL17 symptom scales, treated participants reported significantly higher levels of pain than treatment naïve (Figure 3: Z = −2.538, p = 0.011, *r* = −0.230). Similar to Figure 2, fatigue and insomnia were more prevalent, with the addition of worries and fears related to health and functioning being the most prevalent and poor physical condition being reported more frequently than other symptoms. We observed trends towards a lower physical condition in treated participants (Figure 3: Z = −1.030, p = 0.075, *r* = −0.090) while treatment naïve exhibited trends towards higher worries and fears (Figure 3: Z = −1.680, p = 0.093, *r* = −0.150).

**Figure 3:**
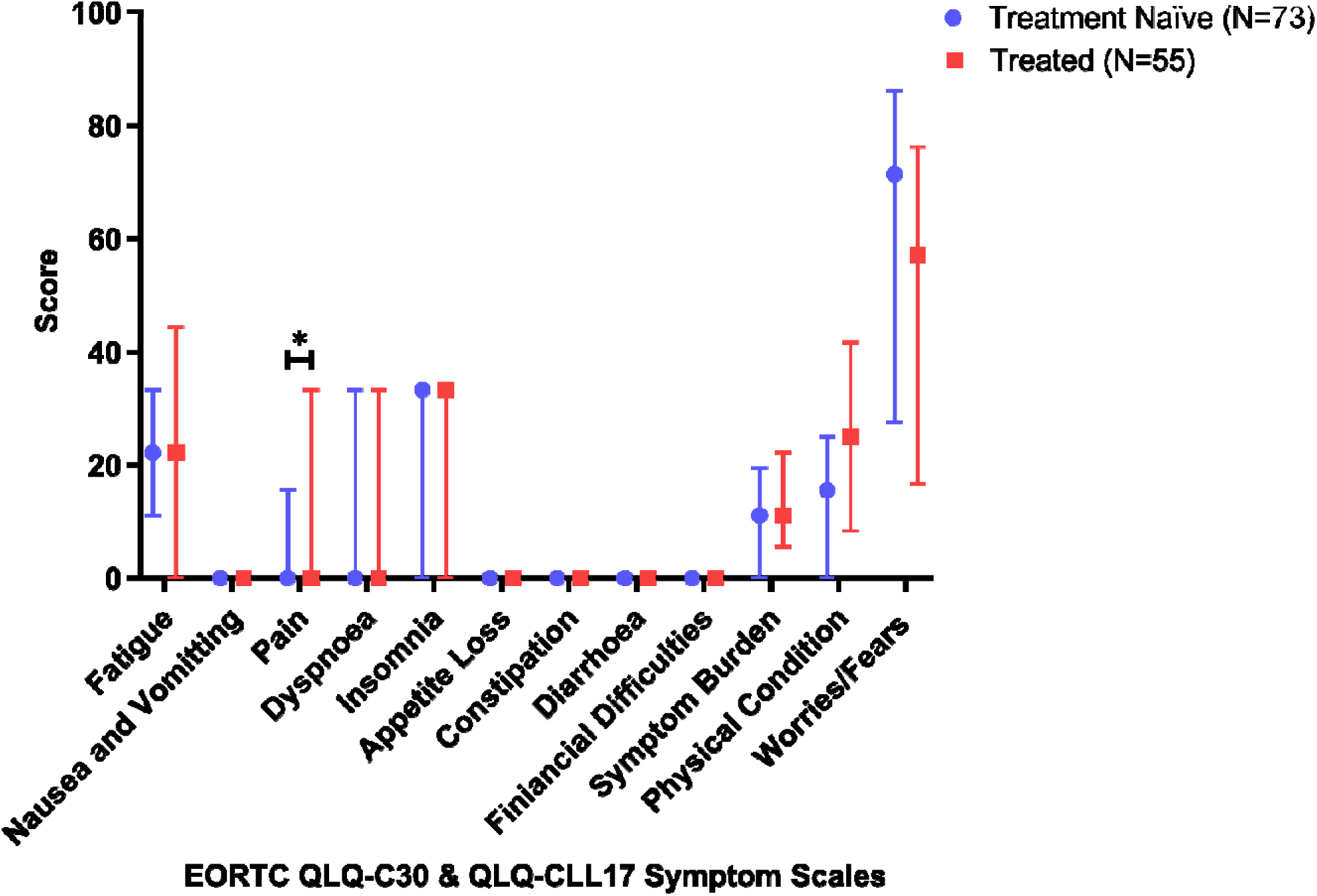
Comparison of symptom scales for quality of life in CLL participants using EORTC QLQ-C30 v3 and EORTC QLQ-CLL17 Questionnaire with treatment naïve compared to those who are treated. All the scales range in scores from 0 to 100; a high score represents a higher (worse) level of symptoms. Values are median and interquartile ranges. *p-value < 0.05, **p-value < 0.01

### Impact of Treatment Status on Physical Activity Levels

The self-reported physical activity levels of the cohort suggest that ∼70% of participants complete exercise at moderate to hard intensities and amounts (Figure 4). However, when analysing the number of minutes spent in moderate and vigorous activities from the Godin LTPAQ, only 24.6% of individuals meet physical activity guidelines of >150 minutes of moderate-intensity activity and/or >75 minutes a week of vigorous-intensity activity. There were no significant differences between those who met physical activity guidelines before diagnosis (29.9%) and those currently meeting physical activity guidelines (24.6%) (Figure 4A: p=0.210). When categorised into activity levels, current physical activity levels remained similar to physical activity before diagnosis (Figure 4A: p = 0.555). However, when categorised further into treatment groups, there was a trend for differences between physical activity levels before and current physical activity levels of those who are/have been treated with more treated people becoming less active (Figure 4A: p=0.072). We observed significant differences between treatment groups and physical activity levels (Figure 4B: p=0.007), which was characterised by 26.5% more treated participants being insufficiently active (Figure 4B: p=0.002) and 22.3% more treatment naïve participants being active (Figure 4B: p=0.016) based on pairwise chi-squared tests with a Bonferroni correction significance threshold of α=0.0167. Additionally, treatment naïve individuals completed around twice as many moderate & vigorous minutes per week after diagnosis compared to treated participants (Treatment Naïve: median = 30.0 (IQR: 20.0-43.0) vs Treated: median = 15.0 (IQR: 5.0-46.5), p=0.045, Figure 4C).

**Figure 4:**
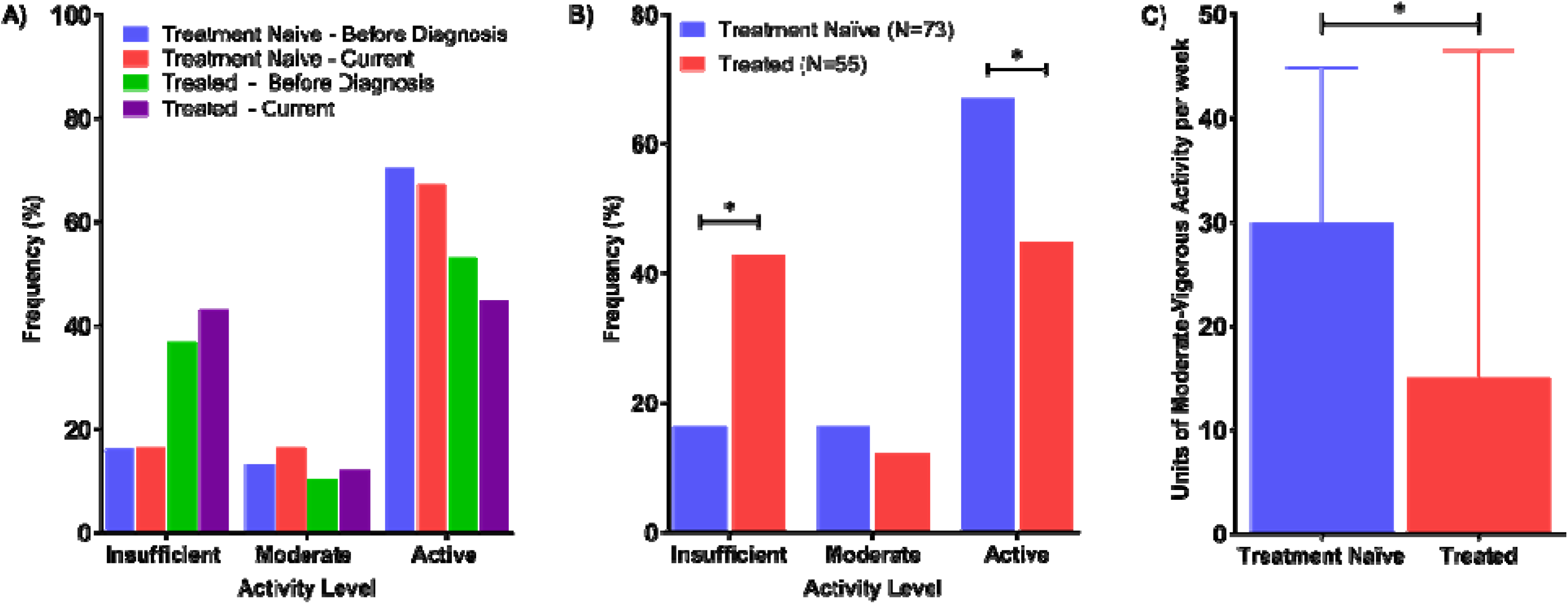
(A) Before diagnosis and current physical activity levels of Treatment Naïve and Treated CLL participants using Godin Leisure Time Physical Activity Questionnaire. (B) Physical activity levels of CLL participants between treatment groups (Treatment Naïve vs Treated) using Godin Leisure Time Physical Activity Questionnaire. (C) Moderate-vigorous physical activity units per week between treatment groups using the Godin Leisure Time Physical Activity Questionnaire. *p-value < 0.05, **p-value < 0.01

### Impact of Physical Activity Levels on QoL and Comorbidities

Based on the analysis of responses to the Godin LTPAQ, post hoc analysis (Dunn’s pairwise test) indicates that CLL participants with insufficient physical activity levels exhibit lower global health status and functional scales than those who are sufficiently active (Figure 5A). Specifically, physically inactive participants have the lowest global health status, a key indicator of QoL (p = 0.020) and the lowest role functioning (p = 0.020) compared to their more active counterparts. Moreover, participants engaging in insufficient (p = 0.003) or moderate (p = 0.042) levels experienced lower physical functioning than the most active.

**Figure 5:**
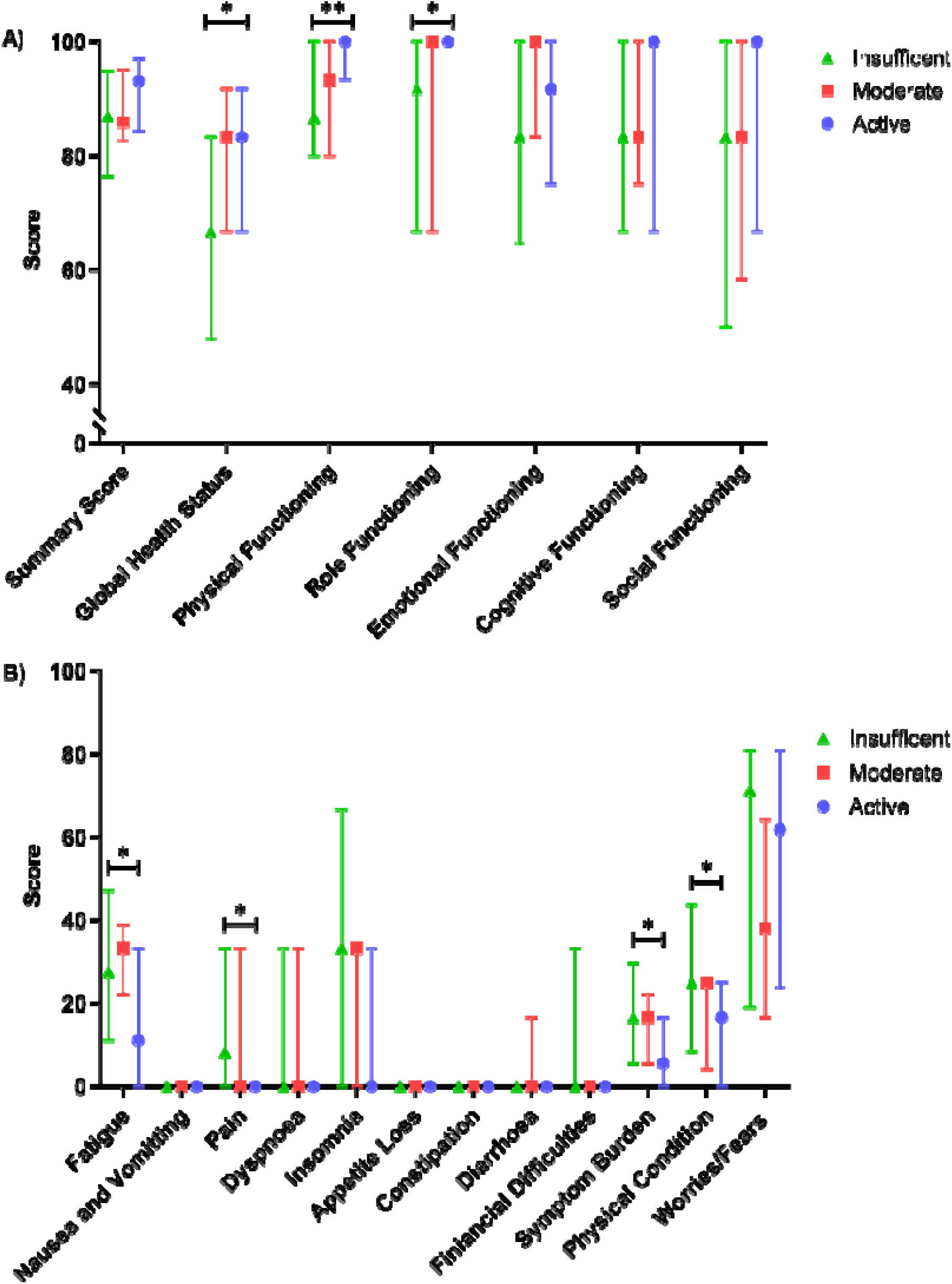
(A) Comparison of global health status and functional scales of quality of life in CLL participants with different physical activity levels categorised based on the Godin Leisure Time Physical Activity Questionnaire. (B) Comparison of symptom scales in quality of life in CLL participants with different physical activity Levels based on the Godin Leisure Time Physical Activity Questionnaire. All the scales range from 0 to 100; a high score represents a higher (better) level of functioning or a higher (worse) level of symptoms. Values are median and interquartile ranges. *p-value < 0.05, **p-value < 0.01

CLL participants with insufficient physical activity levels reported significantly higher symptom severity than sufficiently active participants in several categories (Figure 5B). Insufficiently active participants reported significantly higher levels of fatigue (p = 0.036), pain (p = 0.017), overall symptom burden (p = 0.026), and a decline in physical condition (p = 0.023) compared to those classified as active after post hoc analysis. However, there were no significant differences in these symptoms between insufficiently- and moderately-active participants, nor between moderately- and highly-active participants. When participants were categorised according to the SBAS, no significant differences were observed between the physical activity groups on either the functional or symptomatic scales (data not shown). Similarly, no significant differences in comorbidity scores were found between the physical activity groups, as measured by the Godin LTPAQ (data not shown).

### Associations with Symptoms, QoL and Being More Physically Active

We completed a multivariable linear regression analysis to determine the relationships between clinical factors, symptoms, QoL and whether people met physical activity guidelines (Table 2). Our analysis reveals that CLL treatment, fatigue [OR 0.979; 95% CI (0.960, 0.998), p=0.033], dyspnoea [OR 0.978; 95% CI 0.958, 0.999, p=0.036], insomnia [OR 0.983; 95% CI (0.969, 0.997), p=0.017], physical condition [OR 0.967; 95% CI (0.945, 0.990), p=0.005], and symptom burden [OR 0.960; 95% CI (0.929, 0.992), p=0.014] were associated with a reduced likelihood of meeting physical activity guidelines. Additionally, a trend was observed for pain [OR 0.981; 95% CI (0.962, 1.001), p=0.066] associated with a reduced likelihood. Furthermore, QoL as measured by global health status [OR 1.029; 95% CI (1.007, 1.051), p=0.008] and physical functioning [OR 1.059; 95% CI (1.015, 1.105), p=0.008] were associated with the likelihood of meeting physical activity guidelines while role [OR 1.015; 95% CI (0.997, 1.034), p=0.099] and emotional [OR 1.012; 95% CI (0.999, 1.037), p=0.069] functioning and the Summary Score [OR: 1.038; 95% CI (0.996,1.081), p=0.074] trended towards the likelihood.

**Table 2.**
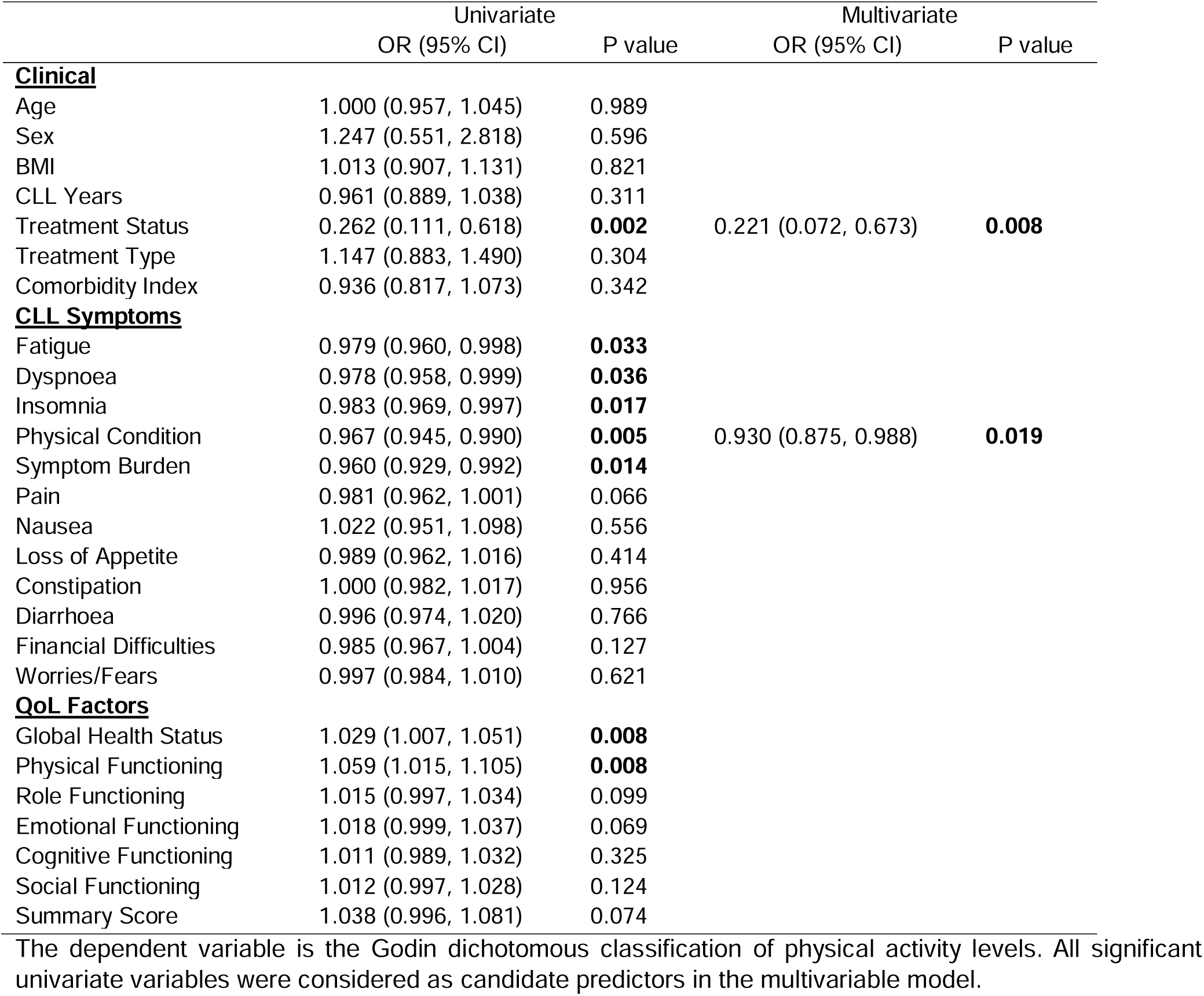
Univariable and multivariable logistic regression models to assess relationships with CLL symptoms and QoL factors and being physically active.

|  | Univariate |  | Multivariate |  |
| --- | --- | --- | --- | --- |
|  | OR (95% CI) | P value | OR (95% CI) | P value |
| **<u>Clinical</u>** |  |  |  |  |
| Age | 1.000 (0.957, 1.045) | 0.989 |  |  |
| Sex | 1.247 (0.551, 2.818) | 0.596 |  |  |
| BMI | 1.013 (0.907, 1.131) | 0.821 |  |  |
| CLL Years | 0.961 (0.889, 1.038) | 0.311 |  |  |
| Treatment Status | 0.262 (0.111, 0.618) | **0.002** | 0.221 (0.072, 0.673) | **0.008** |
| Treatment Type | 1.147 (0.883, 1.490) | 0.304 |  |  |
| Comorbidity Index | 0.936 (0.817, 1.073) | 0.342 |  |  |
| **<u>CLL Symptoms</u>** |  |  |  |  |
| Fatigue | 0.979 (0.960, 0.998) | **0.033** |  |  |
| Dyspnoea | 0.978 (0.958, 0.999) | **0.036** |  |  |
| Insomnia | 0.983 (0.969, 0.997) | **0.017** |  |  |
| Physical Condition | 0.967 (0.945, 0.990) | **0.005** | 0.930 (0.875, 0.988) | **0.019** |
| Symptom Burden | 0.960 (0.929, 0.992) | **0.014** |  |  |
| Pain | 0.981 (0.962, 1.001) | 0.066 |  |  |
| Nausea | 1.022 (0.951, 1.098) | 0.556 |  |  |
| Loss of Appetite | 0.989 (0.962, 1.016) | 0.414 |  |  |
| Constipation | 1.000 (0.982, 1.017) | 0.956 |  |  |
| Diarrhoea | 0.996 (0.974, 1.020) | 0.766 |  |  |
| Financial Difficulties | 0.985 (0.967, 1.004) | 0.127 |  |  |
| Worries/Fears | 0.997 (0.984, 1.010) | 0.621 |  |  |
| **<u>QoL Factors</u>** |  |  |  |  |
| Global Health Status | 1.029 (1.007, 1.051) | **0.008** |  |  |
| Physical Functioning | 1.059 (1.015, 1.105) | **0.008** |  |  |
| Role Functioning | 1.015 (0.997, 1.034) | 0.099 |  |  |
| Emotional Functioning | 1.018 (0.999, 1.037) | 0.069 |  |  |
| Cognitive Functioning | 1.011 (0.989, 1.032) | 0.325 |  |  |
| Social Functioning | 1.012 (0.997, 1.028) | 0.124 |  |  |
| Summary Score | 1.038 (0.996, 1.081) | 0.074 |  |  |

The dependent variable is the Godin dichotomous classification of physical activity levels. All significant univariate variables were considered as candidate predictors in the multivariable model.

Since individual factors associated with physical activity were strongly related to each other, many were not independently predictive in a multivariate stepwise mode. Both treatment status (OR 0.221; 95% CI 0.072, 0.0673, p=0.008) and physical condition [OR 0.930; 95% CI (0.875, 0.988), p=0.019] were independently associated with physical activity.

Given that physical activity levels fluctuate, and some people hover between meeting and achieving recommendations, we determined the relationships between symptoms and the amount of physical activity using a multivariable linear regression analysis of all the symptoms and amounts of physical activity. Symptom burden [β = −0.214 (95% CI: −0.833, −0.083), p=0.017] independently explained 4.6% of the variation in physical activity scores [F(1,122) = 5.85, p=0.017]. As symptom burden is a construct of several different symptoms, we ran a second multivariable analysis to determine which symptoms were associated with symptom burden. We found that fatigue [β = 0.413 (95% CI: 0.155, 0.337), p<0.001], insomnia [β =0.270 (95% CI: 0.061, 0.179), p<0.001] and pain [β =0.271 (95% CI: 0.078, 0.265), p<0.001] independently explained 58.2% of the variation in symptom burden [F(13,114) = 52.84, p<0.001] after Bonferroni correction.

### Exercise Programme Preferences

Most participants (70.3%) reported never receiving guidance regarding exercise or maintaining physical activity following a CLL diagnosis from healthcare professionals (Figure 6). Despite this lack of guidance, 78.9% of respondents expressed interest in participating in an exercise program. Among those who were not interested (N=30), 63.3% believed they were already sufficiently physically active and, therefore, not interested in a programme. Additional reasons for lack of interest included the perception that the program needed to be explicitly tailored to CLL (10%), a preference for exercising alone (10%) and concerns about travel distance (6.6%).

**Figure 6:**
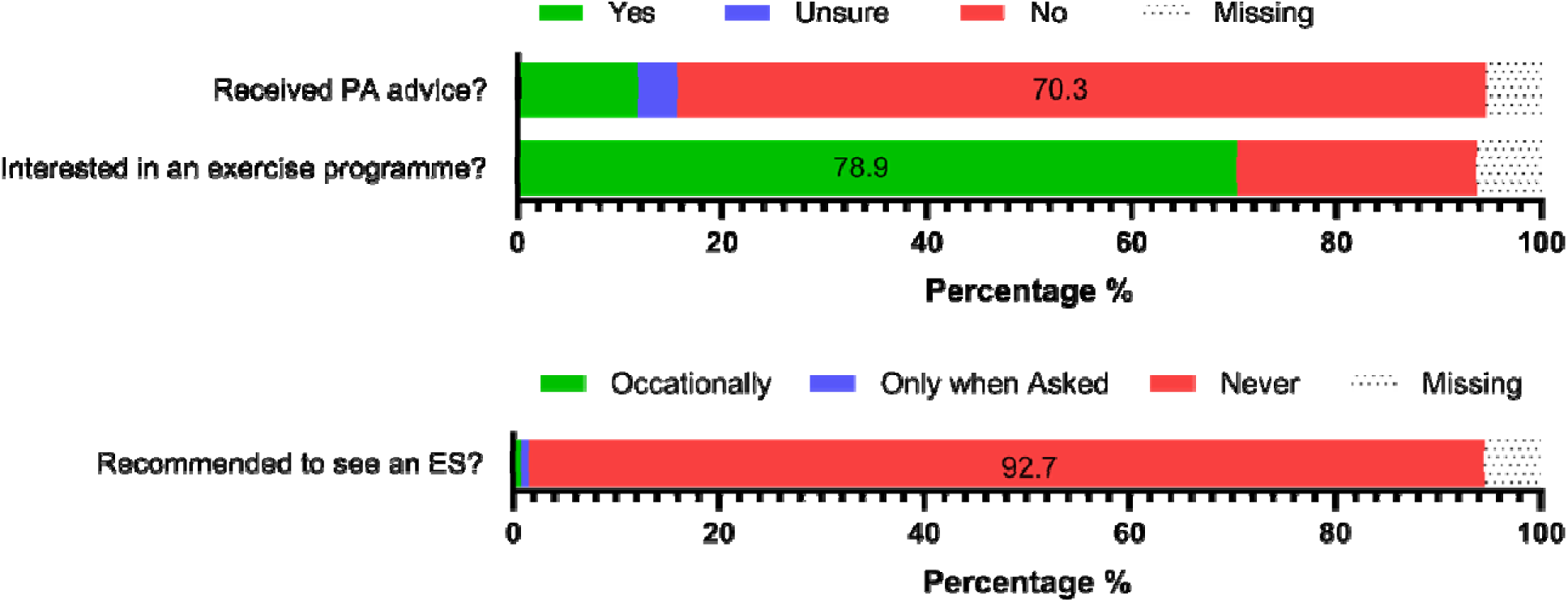
Reported physical activity advice received from health care professionals and respondents interested in an exercise programme. PA, Physical Activity; ES, Exercise Specialist.

The survey also explored preferences regarding the treatment stage, location, structure, and delivery of exercise programs (Figure 7). Participants strongly preferred supervised exercise sessions led by professionals specialising in cancer care. Specifically, 70.3% favoured exercise physiologists, and 58.6% preferred physiotherapists with cancer expertise, whereas only 26.6% agreed with sessions led by personal trainers and 14.8% with sessions conducted by fellow CLL participants. Additionally, respondents favoured exercise programs that included a social component, either with other cancer participants (32.8% in agreement, 35.9% neutral) or with CLL participants and their carers (46.9% in agreement, 31.3% neutral), over one-on-one sessions (25.8% in agreement, 40.6% neutral). The least favoured option was exercising in groups with the general public (19.5% in agreement). Participants expressed a clear preference for exercise programs that offered flexible scheduling (77.3%), were low-cost (70.3%), had consistently available parking (66.4%), were located close to home (72.7%), had good transport links (59.4%), and adhered to robust COVID-19 safety protocols (58.6%). While there were some preferences for virtual exercise programs that could be conducted at home (50.8% in agreement, 25.8% neutral), there were also preferences for community-based clinic programs (43.8% in agreement, 35.9% neutral) and sessions held in private practices with physiotherapists or exercise physiologists (34.4% in agreement, 39.8% neutral). In contrast, conducting exercise programs at the hospital where participants were diagnosed or treated was less popular (19.5% in agreement).

**Figure 7:**
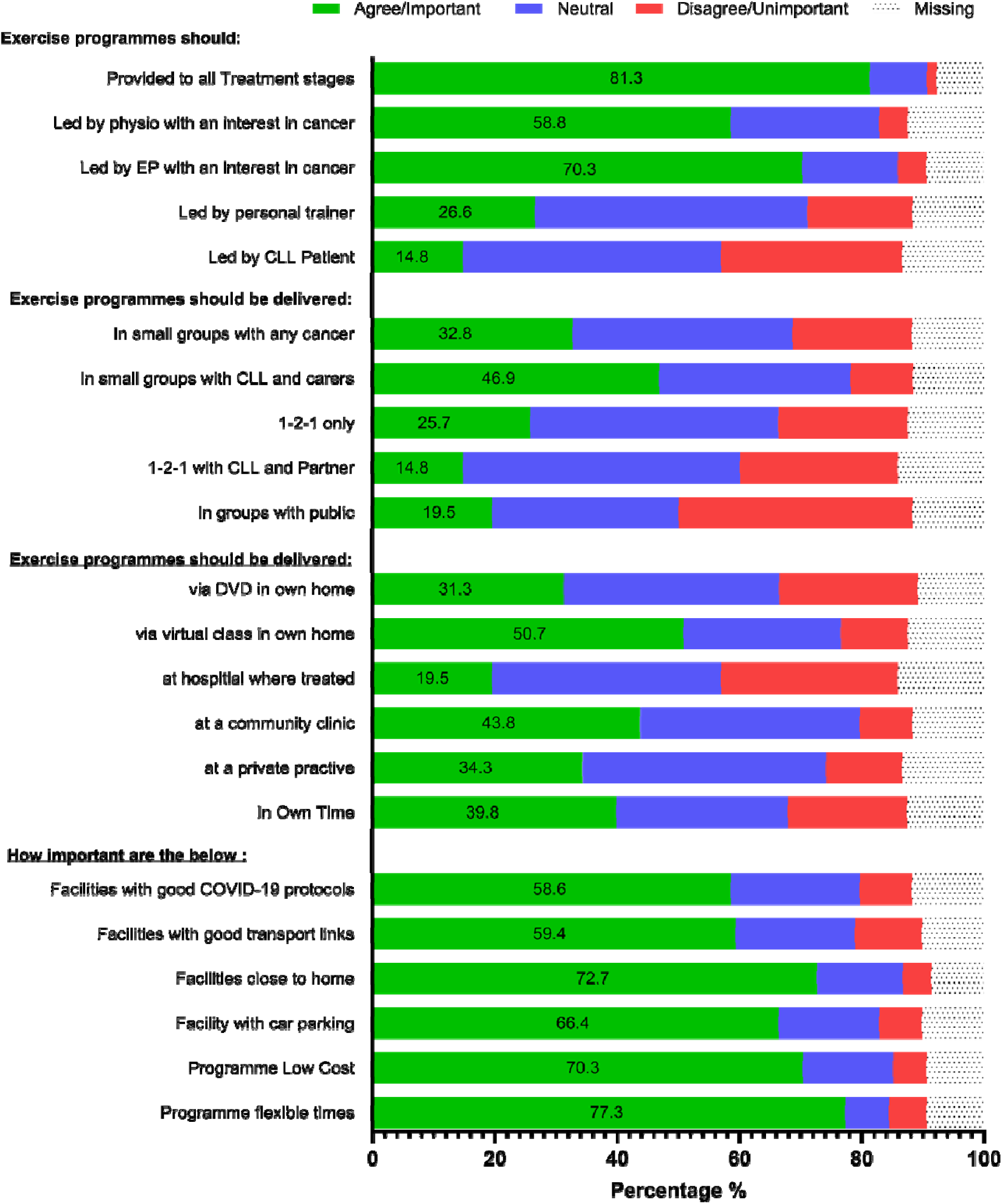
Preference of respondents for treatment stage, location, composition and delivery of exercise programme. EP, Exercise Physiologist.

## Discussion

The relationships between CLL and a reduced QoL are well documented; however, there remains limited information about how potential lifestyle approaches can offset or improve QoL. We report the findings from a UK-based CLL survey that evaluates QoL, symptoms, treatment status and their relationships with physical activity levels. As expected, treatment of CLL was associated with worse QoL indices, while fatigue, insomnia and stress/worries/fears were the most commonly reported symptoms in both treatment groups. Although our cohort self-reported a relatively high engagement in moderate-to vigorous-intensity physical activity, less than one-quarter met the recommended amounts of physical activity per week. People who had not received treatment for CLL engaged in more moderate-to vigorous-intensity physical activity per week and reported a better QoL but had similar symptomology to those who had received treatment. Physically active individuals had lower fatigue, pain, and symptom burdens and a better QoL, as measured by global health status. In multivariate analyses, we found that treatment status and physical condition were independently associated with completing physical activity guidelines. Furthermore, symptom burden, a construct of fatigue, insomnia and pain, was independently associated with the amount of physical activity completed. Our data suggests that engagement with physical activity could help improve QoL by reducing symptoms, which could lower fatigue. However, we highlight a lack of advice about physical activity and exercise participation even though most people (especially those who are inactive) would like specialised physical activity advice and programs from their healthcare professionals.

### Treatment Status, QoL and Physical Activity (QoL v Symptoms)

Our findings indicate that people with CLL who are treatment naïve experienced a better QoL. Specifically, they had better global health status, social functioning and trends towards better role functioning, similar to previous studies in cohorts from other countries (6, 8, 33, 34). Additionally, both treatment groups reported low cognitive functioning scores. Although we did not assess healthy age-matched controls, we can assume and compare from previous studies that even our treatment naïve group scores are lower than the general population. Several disease-related factors have been suggested to contribute to this impairment in QoL (6, 8, 33, 34). Unlike other low-grade lymphomas, CLL is associated with increased opportunistic infections caught primarily through exposure during social and family situations. Such infections are associated with significantly higher chances of hospitalisation with patients who are on immune-suppressing targeted therapies at even higher risks (35). As such, the treated group with lower social functioning and a trend towards lower role functioning are likely due to such risks associated with CLL and exposure to environmental factors. This is consistent with literature indicating that cancer treatments can disrupt social relationships and reduce participation in social activities, leading to feelings of isolation, particularly among haematological malignancies (36). Interestingly, while the treated group reported poorer global health status and social function, both groups expressed high levels of worry related to health and functioning, suggesting that psychological distress is a common experience among people with CLL regardless of treatment status (8, 34). In the treatment naïve group, this anxiety could stem from the uncertainty surrounding disease progression, while the treated group likely worry about treatment-related side effects, recurrence and risk of transformation.

Considering this, we show that CLL treatment is associated with completing more light and less hard physical activity and less likelihood of achieving physical activity guidelines. Additionally, being more physically active was associated with a better QoL. Specifically, higher physical activity levels were associated with higher global health status and better physical and role functioning. Our findings support those of others, where physical activity and exercise training can improve several aspects of QoL in solid malignancies, lymphoma and CLL (10, 12, 16, 18). Physical and role functioning are obvious targets of physical activity and exercise, whereby active people are more inclined to complete basic, instrumental and everyday living tasks. Physical activity in older age is usually associated with more social interactions and may teach people with CLL how to integrate their disease with society. Although not significant, we observed that more active people also had higher cognitive and social functioning scores, similar to others (37). As such, healthcare professionals may underappreciate the risks and benefits of physical activity. When patients are put on active monitoring, they feel that ‘nothing is being done’ and those on treatment are advised to avoid areas of infection risk (38). As such, patients can lose the sense of control over their disease, which undoubtedly reduces their QoL. It is clear that being physically active and engaging in regular exercise is associated with enhanced immune function and reduced risks of opportunistic infections and cancer occurrence (39). In CLL, only 12 weeks of exercise training has the potential to improve immune function and QoL (16, 40). Although it is not surprising that physical activity and exercise improves the QoL, CLL also causes several important disease-related symptoms that may further prevent physical activity engagement.

### Treatment Status, Symptoms and Physical Activity

In both our groups, fatigue, insomnia, stress, anxiety and infections were highly reported. Like others who have analysed symptoms of CLL, fatigue was the most reported symptom, and those who were treated for CLL reported more severe fatigue (34). However, the more active people were, the lower their fatigue levels were. Fatigue levels in several cancers have been shown to improve following exercise and physical activity interventions, albeit the mechanisms remain unknown (41). Our regression analyses show that higher CLL symptom burden, primarily a construct of fatigue, insomnia and pain, was associated with less likelihood of being physically active.

Additionally, symptoms such as poor physical condition were independently associated with not completing physical activity guidelines. Taken together, our data suggests that targeting CLL symptoms has the potential to reduce fatigue and improve their QoL. This is consistent with studies that target the constructs of ageing conditions such as frailty (14), which is also highly prevalent in CLL. By targeting specific symptoms rather than generalised exercise/physical activity, exercise appears as if it can be utilised as a ‘medicinal’ intervention that not only treats symptoms but alleviates factors reducing QoL (18, 42). As such, given that the more active participants reported lower symptom burden and better physical condition, physical activity likely plays a prominent role in CLL symptom management. In other cancers, particularly pre-surgical cancers, multidisciplinary approaches, including exercise, diet and well-being, are utilised to enhance physical function and limit cancer side effects (43). That said, fatigue is clearly associated with the pathophysiology of CLL, as evidenced by high fatigue levels in treatment naïve patients. Therefore, although physical activity-based approaches could reduce fatigue, in some cases (i.e., higher disease burden), the disease pathology may prevent improvements, and maintenance would be considered a positive response. However, such approaches are not offered for people with CLL but are becoming more common in stem cell transplant, showing improved physical condition and fitness (40). Therefore, to investigate whether people with CLL would want such approaches, we asked them about their experiences and preferences.

### Physical Activity Preferences

Similar to patients with multiple myeloma, nearly 80% of our participants suggested they would like to participate in exercise and physical activity programmes (24). However, with 70% having never received guidance from their healthcare professionals and 93% never being recommended to see an exercise specialist, this is a complex conundrum. Not only are most people with CLL older, but they are also immunocompromised, which increases anxiety about safety. Indeed, safety appears to be a primary reason healthcare professionals do not engage in physical activity discussions and referrals (44). Considering that exercise has been shown to be safe, feasible and provide powerful health benefits in CLL and other haematologic malignancies, the lack of guidance is concerning (16–18, 45). However, given that the majority of our participants would not want to exercise at the hospital they are treated at and the lack of facilities in many hospitals, it is understandable that healthcare professionals do not know how to engage patients.

Indeed, most people would want to exercise in a virtual class at home or at a community clinic, not in public groups. Critically, cost, flexibility, and distance from home were most desired if sessions were outside of their home. As such, participants agreed more with exercise programs supervised by cancer care specialists, such as exercise physiologists and physiotherapists. Only a small percentage were interested in programs led by fellow CLL participants, and just 26.6% supported guidance from personal trainers. This suggests that participants place a high value on clinical expertise rather than on peer-led or general fitness instruction, possibly due to the complex nature of their condition, just not in a hospital environment. Taken together, our survey indicates a desire for patients with CLL to be provided with exercise programs at all stages of their disease and to be included as part of clinically focused care. At present, there are no plans for patients with CLL to receive prehabilitation or rehabilitation programs on the NHS; as far as we can determine, no countries provide these services.

### Strengths & Limitations

Strengths of our survey include a good representation of people with CLL in the UK, including treatment conditions, all disease stages, ages and respondents from all education levels. There is always a tendency for younger people who engage in physical activity to respond more to these surveys. However, this was not apparent in our survey. Other strengths included using validated questionnaires that distinguish between active and inactive older populations and have been used to assess physical activity levels in patients with cancer (20, 21, 23). Secondly, recruitment through a CLL charity database is a strength. The database consists of almost 10% of the total number of people with CLL in the UK, providing enough power to accurately represent the CLL population. That said, a potential limitation of our study is that our participants were highly educated. Although similar numbers to the general population hold an undergraduate degree, almost 40% of our group hold postgraduate qualifications. While lower-educated people can be less physically active and have more comorbidities, this was not evident in our cohort. That said, including more from this demographic would likely only have strengthened our results; however, future work should focus on including people from different demographic backgrounds to confirm this. Other limitations include relying on self-reported questionnaires to evaluate physical activity levels, which introduces vulnerability to recall and social desirability biases, especially when relying on retrospective assessments before diagnosis. This could have led to over-or underestimating levels. Using an online survey to collect data poses a limitation, particularly in older individuals and may have contributed to the skewing of education levels. This population may have limited access to or comfort with digital technology, potentially excluding less digitally literate participants and skewing the sample toward those more familiar with online platforms. Finally, although we asked people about their blood counts, including the prevalence of anaemia, less than 10% of participants knew their values or responded to the questions. Given that we have shown previous relations between physical dysfunction and blood markers, this could have added more depth to our understanding.

### Conclusion

Our findings highlight the importance of and desire for targeted interventions to increase physical activity to potentially improve QoL by reducing symptoms, such as fatigue, in individuals with CLL. Our findings suggest that CLL care would likely improve by including exercise advice and prescriptions from healthcare professionals. Currently, most physical activity and exercise approaches employ a ‘cookie-cutter’ approach and try to fit everyone into one model. We show that several factors, including treatment status and symptomology, should be considered when creating CLL-specific programs. Future work is needed to understand the optimal amounts and timing of interventions and the degree of multimodality (e.g., nutrition, well-being and exercise) required while also exploring patient preferences for exercise programmes further to improve accessibility and adherence. Moreover, further research is warranted to clarify the bidirectional relationship between fatigue and physical activity, which could inform more effective, personalised interventions.

## Author Contributions

Conceptualisation: DBB, EEM, RW, SI, EDH, AS.; Data Collection and Management: EEM, HF, AR, ATH, CJ, CLLSA; Data Analysis: EEM, EDH, DBB; Data Interpretation: EEM, JLN, ATH, CJ, RW, SI, EDH, AS, DBB. First draft was written by EEM and all authors commented on previous versions of the manuscript. All uthors critically reviewed the manuscript a nd approved the final version.

## Data Availability

All data produced in the present study are available upon reasonable request to the authors

## References

1. Cancer Research UK. Chronic lymphocytic leukaemia (CLL) statistics 2022 [

2. Hallek M, Al-Sawaf O. Chronic lymphocytic leukemia: 2022 update on diagnostic and therapeutic procedures. Am J Hematol. 2021;96(12):1679-705.

3. Duchesneau ED, McNeill AM, Schary W, Pate V, Lund JL. Prognosis of older adults with chronic lymphocytic leukemia: A Surveillance, Epidemiology, and End Results-Medicare cohort study. Journal of Geriatric Oncology. 2023;14(8):101602.

4. Goede V, Bahlo J, Chataline V, Eichhorst B, Dürig J, Stilgenbauer S, et al. Evaluation of geriatric assessment in patients with chronic lymphocytic leukemia: Results of the CLL9 trial of the German CLL study group. Leuk Lymphoma. 2016;57(4):789–96.

5. Sitlinger A, Thompson DP, Deal MA, Garcia E, Stewart T, Guadalupe E, et al. Exercise and chronic lymphocytic leukemia (CLL)-relationships among physical activity, fitness, & inflammation, and their impacts on CLL patients. Blood. 2018;132:5540.

6. Holzner B, Kemmler G, Kopp M, Nguyen-Van-Tam D, Sperner-Unterweger B, Greil R. Quality of life of patients with chronic lymphocytic leukemia: results of a longitudinal investigation over 1 yr. Eur J Haematol. 2004;72(6):381–9.

7. Shanafelt TD, Bowen DA, Venkat C, Slager SL, Zent CS, Kay NE, et al. The physician-patient relationship and quality of life: lessons from chronic lymphocytic leukemia. Leuk Res. 2009;33(2):263–70.

8. Youron P, Singh C, Jindal N, Malhotra P, Khadwal A, Jain A, et al. Quality of life in patients of chronic lymphocytic leukemia using the EORTC QLQ-C30 and QLQ-CLL17 questionnaire. European journal of haematology. 2020;105(6):755–62.

9. Fedele PL, Opat S. Chronic Lymphocytic Leukemia: Time to Care for the Survivors. Journal of Clinical Oncology. 2024;42(17):2005–11.

10. Campbell KL, Winters-Stone KM, Wiskemann J, May AM, Schwartz AL, Courneya KS, et al. Exercise Guidelines for Cancer Survivors: Consensus Statement from International Multidisciplinary Roundtable. Med Sci Sports Exerc. 2019;51(11):2375–90.

11. Cristian A, Ruiz M. Comparison of Function, Fatigue, Frailty in Patients with Malignant Hematology, Pre-Bone Marrow Transplant and Solid Tumors. Biology of Blood and Marrow Transplantation. 2020;26(3):S362.

12. Sitlinger A, Brander DM, Bartlett DB. Impact of exercise on the immune system and outcomes in hematologic malignancies. Blood advances. 2020;4(8):1801–11.

13. Ofori-Asenso R, Chin KL, Mazidi M, Zomer E, Ilomaki J, Zullo AR, et al. Global incidence of frailty and prefrailty among community-dwelling older adults: a systematic review and meta-analysis. JAMA network open. 2019;2(8):e198398-e.

14. Taylor JA, Greenhaff PL, Bartlett DB, Jackson TA, Duggal NA, Lord JM. Multisystem physiological perspective of human frailty and its modulation by physical activity. Physiological Reviews. 2023;103(2):1137–91.

15. Bellizzi KM, Rowland JH, Arora NK, Hamilton AS, Miller MF, Aziz NM. Physical activity and quality of life in adult survivors of non-Hodgkin’s lymphoma. Journal of Clinical Oncology. 2009;27(6):960–6.

16. MacDonald G, Sitlinger A, Deal MA, Hanson ED, Ferraro S, Pieper CF, et al. A pilot study of high-intensity interval training in older adults with treatment naïve chronic lymphocytic leukemia. Scientific reports. 2021;11(1):23137.

17. Crane JC, Gordon MJ, Basen-Engquist K, Ferrajoli A, Markofski MM, Lee CY, et al. Relationships between T-lymphocytes and physical function in adults with chronic lymphocytic leukemia: Results from the HEALTH4CLL pilot study. European Journal of Haematology. 2023;110(6):732–42.

18. Artese AL, Sitlinger A, MacDonald G, Deal MA, Hanson ED, Pieper CF, et al. Effects of high-intensity interval training on health-related quality of life in chronic lymphocytic leukemia: A pilot study. Journal of Geriatric Oncology. 2023;14(1):101373.

19. Taylor-Piliae RE, Fair JM, Haskell WL, Varady AN, Iribarren C, Hlatky MA, et al. Validation of the Stanford Brief Activity Survey: examining psychological factors and physical activity levels in older adults. Journal of Physical Activity and Health. 2010;7(1):87–94.

20. Taylor-Piliae RE, Norton LC, Haskell WL, Mahbouda MH, Fair JM, Iribarren C, et al. Validation of a new brief physical activity survey among men and women aged 60–69 years. American journal of epidemiology. 2006;164(6):598–606.

21. Godin G. The Godin-Shephard leisure-time physical activity questionnaire. The Health & Fitness Journal of Canada. 2011;4(1):18–22.

22. Tillmann V, Darlington A, Eiser C, Bishop N, Davies H. Male sex and low physical activity are associated with reduced spine bone mineral density in survivors of childhood acute lymphoblastic leukemia. Journal of bone and mineral research. 2002;17(6):1073–80.

23. Amireault S GG, Lacombe J, Sabiston CM. The use of the Godin-Shephard Leisure-Time Physical Activity Questionnaire in oncology research: a systematic review. BMC medical research methodology. BMC medical research methodology. 2015;15(1):1–11.

24. Nicol JL, Woodrow C, Burton NW, Mollee P, Nicol AJ, Hill MM, et al. Physical Activity in People with Multiple Myeloma: Associated Factors and Exercise Program Preferences. J Clin Med. 2020;9(10).

25. Department of Health and Social Care. UK Chief Medical Officers’ Physical Activity Guidelines. In: Care DoHaS, editor. GOV.UK 2019.

26. Aaronson NK, Ahmedzai S, Bergman B, Bullinger M, Cull A, Duez NJ, et al. The European Organization for Research and Treatment of Cancer QLQ-C30: a quality-of-life instrument for use in international clinical trials in oncology. J Natl Cancer Inst. 1993;85(5):365–76.

27. Fayers P, Bottomley A, Group EQoL. Quality of life research within the EORTC—the EORTC QLQ-C30. European Journal of Cancer. 2002;38:125–33.

28. van de Poll-Franse L, Oerlemans S, Bredart A, Kyriakou C, Sztankay M, Pallua S, et al. International development of four EORTC disease-specific quality of life questionnaires for patients with Hodgkin lymphoma, high- and low-grade non-Hodgkin lymphoma and chronic lymphocytic leukaemia. Qual Life Res. 2018;27(2):333–45.

29. Fayers P, Aaronson N, Bjordal K, Groenvold M, Curran D, Bottomley A. On behalf of the EORTC quality of life group: the EORTC QLQ-C30 scoring manual. Guidelines for assessing Quality of Life in EORTC Clinical Trials. 2001;3.

30. Linn BS, Linn MW, Gurel L. Cumulative illness rating scale. J Am Geriatr Soc. 1968;16(5):622–6.

31. Gordon MJ, Kaempf A, Sitlinger A, Shouse G, Mei M, Brander DM, et al. The Chronic Lymphocytic Leukemia Comorbidity Index (CLL-CI): A Three-Factor Comorbidity Model. Clin Cancer Res. 2021;27(17):4814–24.

32. Cleghorn CL, Harrison RA, Ransley JK, Wilkinson S, Thomas J, Cade JE. Can a dietary quality score derived from a short-form FFQ assess dietary quality in UK adult population surveys? Public health nutrition. 2016;19(16):2915–23.

33. Holtzer-Goor KM, Schaafsma MR, Joosten P, Posthuma EF, Wittebol S, Huijgens PC, et al. Quality of life of patients with chronic lymphocytic leukaemia in the Netherlands: results of a longitudinal multicentre study. Quality of life research: an international journal of quality of life aspects of treatment, care and rehabilitation. 2015;24(12):2895–906.

34. Shanafelt TD, Bowen D, Venkat C, Slager SL, Zent CS, Kay NE, et al. Quality of life in chronic lymphocytic leukemia: an international survey of 1482 patients. British journal of haematology. 2007;139(2):255–64.

35. Hallek M. Chronic lymphocytic leukemia: 2020 update on diagnosis, risk stratification and treatment. Am J Hematol. 2019;94(11):1266-87.

36. Bryant J, Mansfield E, Hall A, Waller A, Boyes A, Jayakody A, et al. The psychosocial outcomes of individuals with hematological cancers: Are we doing enough high quality research, and what is it telling us? Critical Reviews in Oncology/Hematology. 2016;101:21–31.

37. Younan B. Cognitive Functioning Differences Between Physically Active and Sedentary Older Adults. J Alzheimers Dis Rep. 2018;2(1):93–101.

38. Shanafelt TD, Byrd JC, Call TG, Zent CS, Kay NE. Narrative review: initial management of newly diagnosed, early-stage chronic lymphocytic leukemia. Ann Intern Med. 2006;145(6):435–47.

39. Emery A, Moore S, Turner JE, Campbell JP. Reframing How Physical Activity Reduces The Incidence of Clinically-Diagnosed Cancers: Appraising Exercise-Induced Immuno-Modulation As An Integral Mechanism. Front Oncol. 2022;12:788113.

40. Artese A, Winthrop H, Bohannon L, Lew M, Johnson E, MacDonald G, et al. A pilot study to assess the feasibility of a remotely monitored high-intensity interval training program prior to allogeneic hematopoietic stem cell transplantation. PLoS One. 2023.

41. Mustian KM, Sprod LK, Janelsins M, Peppone LJ, Mohile S. Exercise Recommendations for Cancer-Related Fatigue, Cognitive Impairment, Sleep problems, Depression, Pain, Anxiety, and Physical Dysfunction: A Review. Oncol Hematol Rev. 2012;8(2):81–8.

42. Brown JC, Huedo-Medina TB, Pescatello LS, Pescatello SM, Ferrer RA, Johnson BT. Efficacy of exercise interventions in modulating cancer-related fatigue among adult cancer survivors: a meta-analysis. Cancer Epidemiology, Biomarkers & Prevention. 2011;20(1):123–33.

43. Sadlonova M, Katz NB, Jurayj JS, Flores L, Celano CM, von Arnim CAF, et al. Surgical prehabilitation in older and frail individuals: a scoping review. Int Anesthesiol Clin. 2023;61(2):34–46.

44. Haussmann A, Gabrian M, Ungar N, Jooß S, Wiskemann J, Sieverding M, et al. What hinders healthcare professionals in promoting physical activity towards cancer patients? The influencing role of healthcare professionals’ concerns, perceived patient characteristics and perceived structural factors. Eur J Cancer Care (Engl). 2018;27(4):e12853.

45. Lee CY, Gordon MJ, Markofski MM, LaVoy EC, Peterson SK, Li L, et al. Optimization of mHealth behavioral interventions for patients with chronic lymphocytic leukemia: the HEALTH4CLL study. J Cancer Surviv. 2024.

